# Physiology Guided Machine Learning Approach to Predict Short and Long Term Outcomes of Obstructive Sleep Apnea

**DOI:** 10.1101/2024.11.20.24317571

**Authors:** Sajila D. Wickramaratne, Korey Kam, Thomas M. Tolbert, Andrew Varga, Indu Ayappa, David M. Rapoport, Ankit Parekh

**Author notes:** Corresponding Author: Sajila D. Wickramaratne, PhD, Division of Pulmonary, Critical Care and Sleep Medicine, Icahn School of Medicine at Mount Sinai, New York, NY – 10029.

## Abstract

Obstructive Sleep Apnea(OSA) is a chronic condition that affects 1 billion people worldwide. Apnea Hypopnea Index(AHI) is the clinical gold standard to measure the severity of OSA. This study highlights limitations in the apnea-hypopnea index as a predictor for obstructive sleep apnea (OSA) outcomes. Instead, a physiology-guided machine learning (ML) approach was developed using features from ventilatory, hypoxic, and arousal domains, based on polysomnography data from the Sleep Heart Health Study (SHHS). The ML model demonstrated superior predictive performance for all-cause mortality (AUROC-0.93) and daytime sleepiness (AUROC-0.81) compared to AHI. Explainable AI techniques, such as SHAP analysis, provided insights into feature importance, offering a clinically interpretable and scalable tool for OSA outcome prediction.

## Background

Closer to 1 billion individuals worldwide suffer from Obstructive sleep apnea (OSA) ^1^. It has short- and long-term adverse effects, including excessive daytime sleepiness, neurocognitive impairment, hypertension, and cardio-cerebrovascular morbidity ^1,2^. OSA severity is clinically determined by the apnea-hypopnea index (AHI), and several recent studies have highlighted its limitations, including its subjective nature and poor predictive ability for short- and long-term adverse outcomes^2,3^. AHI can be interpreted as a fixed combination of potentially independent ventilatory/hypoxic/arousal domains. Recent studies suggest that metrics that characterize the severity of OSA along each domain are independently better associated with long-term outcomes of OSA^4–6^.

, offering a clinically interpretable and scalable tool for OSA outcome prediction.

The apnea-hypopnea Index (AHI) is inconsistently associated with adverse outcomes of OSA. This index measures the frequency of respiratory incidents, specifically apneas and hypopneas. The severity of OSA is determined by either the frequency of these incidents or their immediate physiological effects, such as oxygen desaturation and awakening from sleep. OSA is recognized as a complex and heterogeneous disorder, and AHI may not be able to capture this heterogeneity^7,8^. The value of AHI is influenced by various physiological factors, technical variables related to the equipment used, scoring rules, and subjective judgments when establishing baselines for oxygen saturation and airflow^9^.

AHI is a poor predictor of adverse outcomes. AHI can be interpreted as a fixed combination of ventilatory, hypoxic, and arousal domains. Previous research done in the field has found that the metrics along the domains can predict adverse outcomes better than AHI^4,5,10,11^. OSA-specific intermittent hypoxemia is known to have adverse effects on cardio metabolic function ^5,12^. The area defined between O2 desaturations during manually marked respiratory events and the baseline of the O2 saturation curve has been shown to be moderately associated with cardiovascular outcomes in two community-based cohorts: cardiovascular mortality increasing with desaturation area^5^. The association between hypoxic burden and cardiovascular mortality was greater than with AHI or other simple measures of hypoxia. Arousal from sleep is a hallmark of OSA, with frequent arousal from sleep being thought to lead to adverse neurocognitive outcomes^6,13^. Arousal intensity quantified via cortical EEG, or autonomic (pulse-rate) changes was moderately associated with all-cause mortality^11^. Sleep EEG measure the quantifying depth of sleep such as K-complex-based metrics and ORP and are found to be modestly associated with daytime sleepiness^14,15^.

In summary, while several metrics have been posited as useful alternatives for assessing OSA severity, there is currently no unified framework combining these metrics across hypoxia, arousal, and ventilation domains. From current research, we do not observe the usage of a wealth of clinical knowledge associated with OSA being effectively used to predict outcomes using state-of-art machine learning models. This disconnect negatively impacts the field. Hence our physiology–guided machine learning approach that is presented in this article will combine the recent developments from both fields to develop models that can be clinically implemented in the future.

Traditional AI/ML classification/regression models do not consider the time to event, which is a very important factor in predicting survival probabilities. Survival analysis is specifically designed to handle data that focuses on the time it takes for an event of interest, such as all-cause mortality^16^. It is critical in scenarios where it is necessary to know the timing and probability of occurrence of specified events (cardiovascular, neurodegeneration etc.). Survival analyses further allow for comparisons of survival experiences between different groups or populations; these are not possible when outcomes are dichotomized, as is done for current AI/ML models. Several studies have thus suggested the use of AI/ML models that model these complex multivariate survival profiles and can generate the time to event based on input features. This analysis is particularly useful in clinical trials, where such an AI/ML model can evaluate the effectiveness of new treatments or interventions. With the recent developments of machine learning, more complex multivariate survival models have enhanced the survival profile generation in predicting disease progression^17,18^ This methodology has the potential to predict disease progression for adverse outcomes of OSA and guide the clinical decision-making in deciding proper treatment methods.

Even though the AI/ML field is developing rapidly, its use in the OSA, better than that done with AHI, needs to be improved, with current research aimed at rudimentary models with a lesser focus on interpretability or clinical implementation. Here, we propose a physiology-guided ML-based approach to combine metrics along the ventilator / hypoxic/arousal domains to predict OSA’s short- and long-term outcomes and determine the most important features when it comes to predicting OSA’s short- and long-term outcomes. Further, we used our physiology-guided ML approach to predict the survival profiles for the subjects and determine how the feature importance changes through the period.

## Materials & Methods

### System Overview

Data: The data used for this study was obtained from the Sleep Heart Health Study (SHHS). The characteristics of the data cohort are shown in Table 1 in supplementary materials. The preliminary analysis was done with data from the Sleep Heart Health Study data cohort, where sleep studies from N=5,804 subjects were initially analyzed, and 3038 with valid good quality data (EEG, SPO2, and Flow signal) were eventually selected for the study.

**Table 1:** Subject Characteristics.

| Subject Characteristics | Dataset |
| --- | --- |
|  | SHHS v1 |
| N | 3084 |
| Age (yrs.) | 63.39±11.18 |
| Sex (M/F) | 1105/969 |
| AHI3A (/hr.) | 19.52±10.39 |
| Arousal Index (/hr.) | 19.25±10.29 |
| SE (%) | 83.59±10.16 |
| Time in Bed (mins) | 435±53.12 |
| Total Sleep Time (mins) | 363.76±60.63 |
| WASO (mins) | 58.30±41.88 |
| N1 (%) | 5.42±3.88 |
| N2 (%) | 56.59±11.46 |
| N3 (%) | 18.01±11.68 |
| REM (%) | 19.97±6.17 |
| Sleepiness(1/0) | 935/2149 |
| All Cause Mortality(1/0) | 642/2376 |

AHI3A: apnea-hypopnea index with 3% desaturation and/or arousal; SE: sleep efficiency; WASO: wake after sleep onset; REM: rapid eye movement

**Table 2:**

| Status | Time Bins |  |  |  |  |  |
| --- | --- | --- | --- | --- | --- | --- |
|  | (0,1000] | (1000,2000] | (2000,3000] | (3000,4000] | (4000,5000] | (5000,6000] |
| Alive | 4372 | 4141 | 3877 | 3581 | 3469 | 3467 |
| Dead | 129 | 360 | 624 | 920 | 1032 | 1034 |

**Table 3:** The Performance Metrics for the Survival Models.

| **Model** | **C-index** | **Integrated Brier Score** | **Mean Time Dependent AUROC** |
| --- | --- | --- | --- |
| **Random Model** | 0.50 | 0.28 | 0.50 |
| **AHI Model** | 0.546 | 0.155 | 0.572 |
| **AHI+ Demo Model** | 0.76 | 0.092 | 0.795 |
| **VB+HB+AB Model**<br>**(3 vars)** | 0.58 | 0.142 | 0.590 |
| **VB (40) +HB (20)**<br>**+AB (10) Model** | 0.632 | 0.102 | 0.65 |
| **VB (40) +HB (20)**<br>**Model** | 0.57 | 0.141 | 0.621 |
| **VB+HB+AB (10**<br>**vars)** | 0.59 | 0.135 | 0.625 |
| **VB+ HB+ AB+ Demo**<br>**(10 vars)** | 0.77 | 0.088 | 0.800 |
| **VB+ HB+ AB+ Demo** | 0.786 | 0.081 | 0.801 |

**Table 4:**
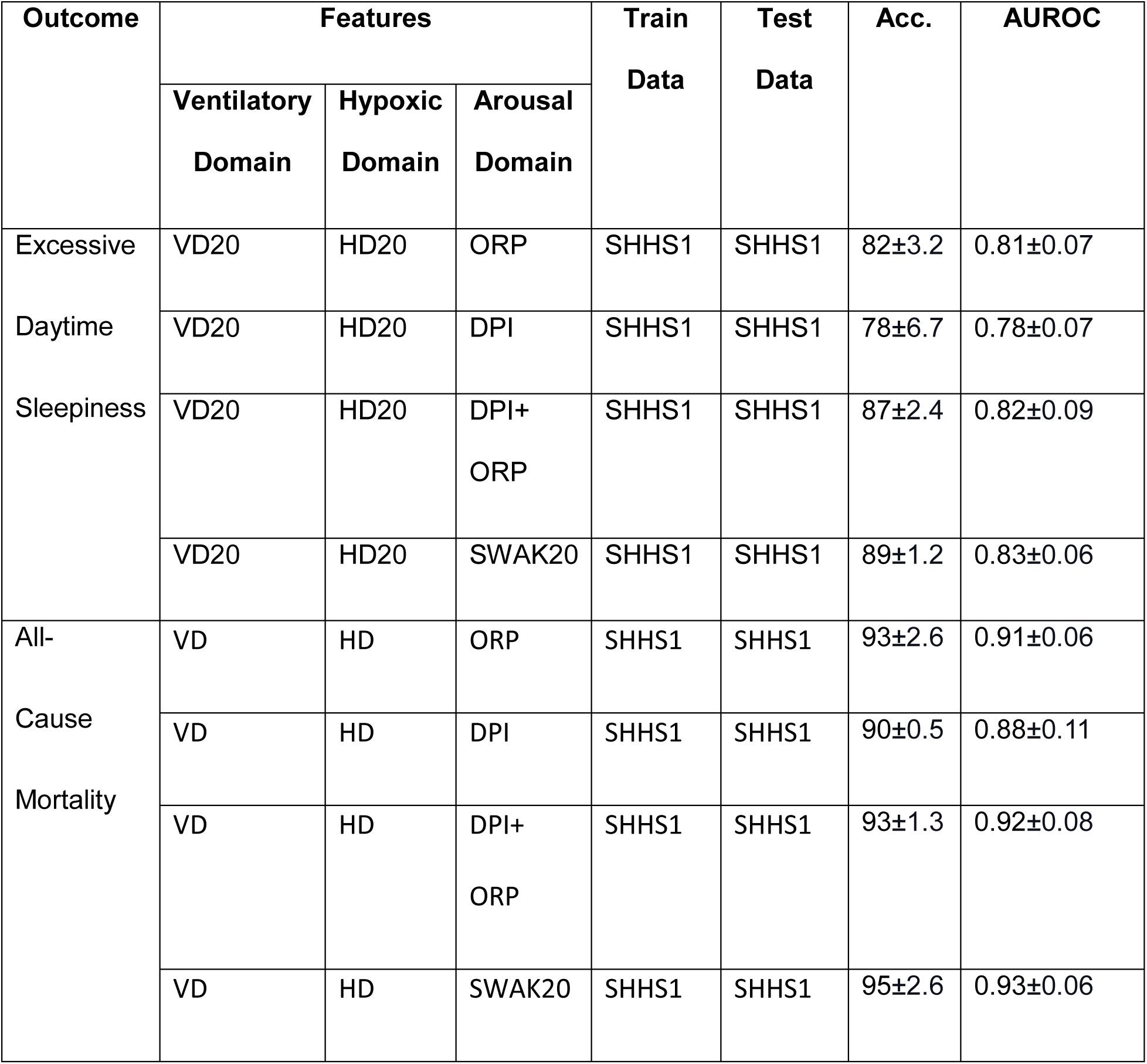
Performance Metrics for the Prediction Models.

To train the machine-learning model features belonging to ventilatory, hypoxic, and arousal domains were extracted as input features from the Flow, SPO2, and EEG channels. For the ventilatory and hypoxic domains, distributions were generated using our automated algorithm. The derivation Arousal domain was represented by Odds Ratio Product (ORP) deciles^14^. The target outcomes considered for this study were Daytime Sleepiness and Risk of All-Cause Mortality. For data from the SHHS, as the nasal cannula was not available, Airflow was derived using a digital differentiator applied to the thoracic and abdominal respiratory inductance plethysmography (RIP) signal.

### Model

The input features used for training the ML models represent VB, HB, and AB. These features represent a whole night of Polysomnography recording. An Extreme Gradient Boosting (XGBoost) model was trained with 70 input features. XGBoost model was chosen after comparing the performance metrics with a pool of ML models. XGBoost is a decision-tree-based ensemble ML algorithm that uses a gradient-boosting framework^19^. XGBoost iteratively builds a set of weak learners on subsets of data to produce a strong learner. It currently dominates in performance when it comes to structured or tabular datasets on classification predictive modeling problems. Due to the tabular nature of our input features, we expect XGBoost to be the best performing ML model for this study. After predicting the outcomes, Shap analysis was performed to determine the feature importance. Shap Analysis is widely used to determine the feature importance of non-linear models, such as XGBoost, used for this study. For comparison, we separately trained a univariate Logistic Regression model with AHI3a as the only input feature for both outcomes.

Our data set had two outcomes: Excessive Daytime Sleepiness and All-Cause Mortality. There was a class imbalance whereby the positive cases for Excessive Daytime Sleepiness and All-Cause Mortality had a significantly lower number of instances compared with negative cases. Techniques such as Synthetic Minority Oversampling Technique (SMOTE) and random under sampling have been widely used in the literature for class imbalance^20^. We applied SMOTE for on all cohorts to achieve a reasonable class balance.

### Excessive Daytime Sleepiness Prediction

The data from SHHS visit 1 was used to develop and test ML models that can predict Daytime Sleepiness. The Daytime Sleepiness was coded using the Epworth Sleepiness Score and 1/0 based on the cutoff score 10 to represent sleepiness and non-sleepiness.

### All-Cause Mortality Prediction

The data from SHHS was used to develop and test ML models that can All-Cause Mortality. Cardiovascular death was also included in All-Cause Mortality data. Mortality and days to the event was confirmed using follow-up interviews, written annual questionnaires, or telephone contacts with study participants or next-of-kin, surveillance of local hospital records and community obituaries, and linkage with the Social Security Administration Death Master File.

### All-Cause Mortality Survival Curve Generation

After outcome prediction using the same input features representing VD, HD, and AD with the addition of time to the event, a survival analysis was performed to forecast the survival curves for the subjects. The gradient-boosting Survival model was used for the survival analysis.

Hyperparameters are values or weights that determine the learning process of an ML algorithm. We can utilize XGBoost efficiently by tuning its large number of hyperparameters. The standard hyperparameter optimization is time-consuming and computationally expensive. Bayesian Optimization is a probabilistic model-based technique we will be utilizing for hyperparameter optimization^21,22^. This approach can yield better performance while requiring fewer iterations by taking into account past evaluations when choosing the optimal set of hyperparameters^23^. We used Bayesian Optimization implemented through the Python libraries HyperOpt^24^ and Optuna Framework^25^.This analysis will use classification and survival models, which require separate tuning mechanisms. For the prediction models, the hyper parameters were tuned to maximize the AUROC value. The hyper parameters of the survival model were tuned to minimize the Integrated Brier Score. The tuned hyper parameters and their values for each model is in the supplementary materials.

### Model Evaluation

All model training and analysis were performed using Python. Normality of data was tested using the Shapiro-Wilk test. Mean and standard deviation are reported for normally distributed variables, whereas median and IQR are reported for non-normally distributed variables.

The Sleepiness and All-Cause Mortality prediction ML models were evaluated using Area Under Operator Receiver Curve (AUROC). The area under the receiver operating characteristics curve (ROC curve) is a popular performance measure for binary classification tasks. Given a predicted risk score the ROC curve compares the false positive rate (1 - specificity) against the true positive rate (sensitivity) for each value of predicted risk score. An AUROC value greater than 0.9 indicates the classifier is excellent in distinguishing between the classes.

Explainable AI SHapley Additive exPlanations (SHAP) is a method to explain predictions. SHAP is based on the game theoretically optimal Shapley values^26^. Tree-based ML models such as gradient-boosted trees are nonlinear predictive models that can benefit from SHAP analysis to understand global and local feature importance and have been used in biomedical applications^27,28^. In this study, SHAP values were used for model interpretation to determine the most important features for predicting each adverse outcome.

The C-index is the most widely used metric for the global evaluation of prognostic models in survival analysis. The C-index is a summary of model performance across a wide array of possible configurations. It’s distinguishing feature, however, is that it can characterize a model’s prognostic ability by accounting for both outcome occurrence and timing (cite). To interpret the C-index value of 0.5 denotes a random model, a value of 1.0 denotes a perfect model and a value of 0.0 denotes a perfectly wrong model.

When extending the ROC curve to continuous outcomes, particularly survival time, a patient’s disease status is typically not fixed and changes over time: at enrollment, a subject is usually healthy but may be diseased later. Consequently, sensitivity and specificity become time-dependent measures.

The time-dependent Brier score is an extension of the mean squared error to right censored data. The Brier score is used to assess calibration and discrimination^29^. For this study all time points between the 10% and 90% percentile of observed time points were used to calculate the integrated Brier Score. Lower Brier scores close to 0 is desirable. Apart from the survival profile generation, the Feature Importance was calculated based on Permutation Importance.

## Results

### All-Cause Mortality Prediction

Fig. 1 shows the ROC curves obtained for predicting the risk of All-Cause Mortality using AHI and our ML model. The ML model obtained an AUROC value of (0.93±0.06), compared to the AHI model with an AUROC of 0.57. All-cause mortality (including cardiovascular death) shows features from all three domains among the most important features, with majority of them coming from the arousal domain.

**Figure 1:**
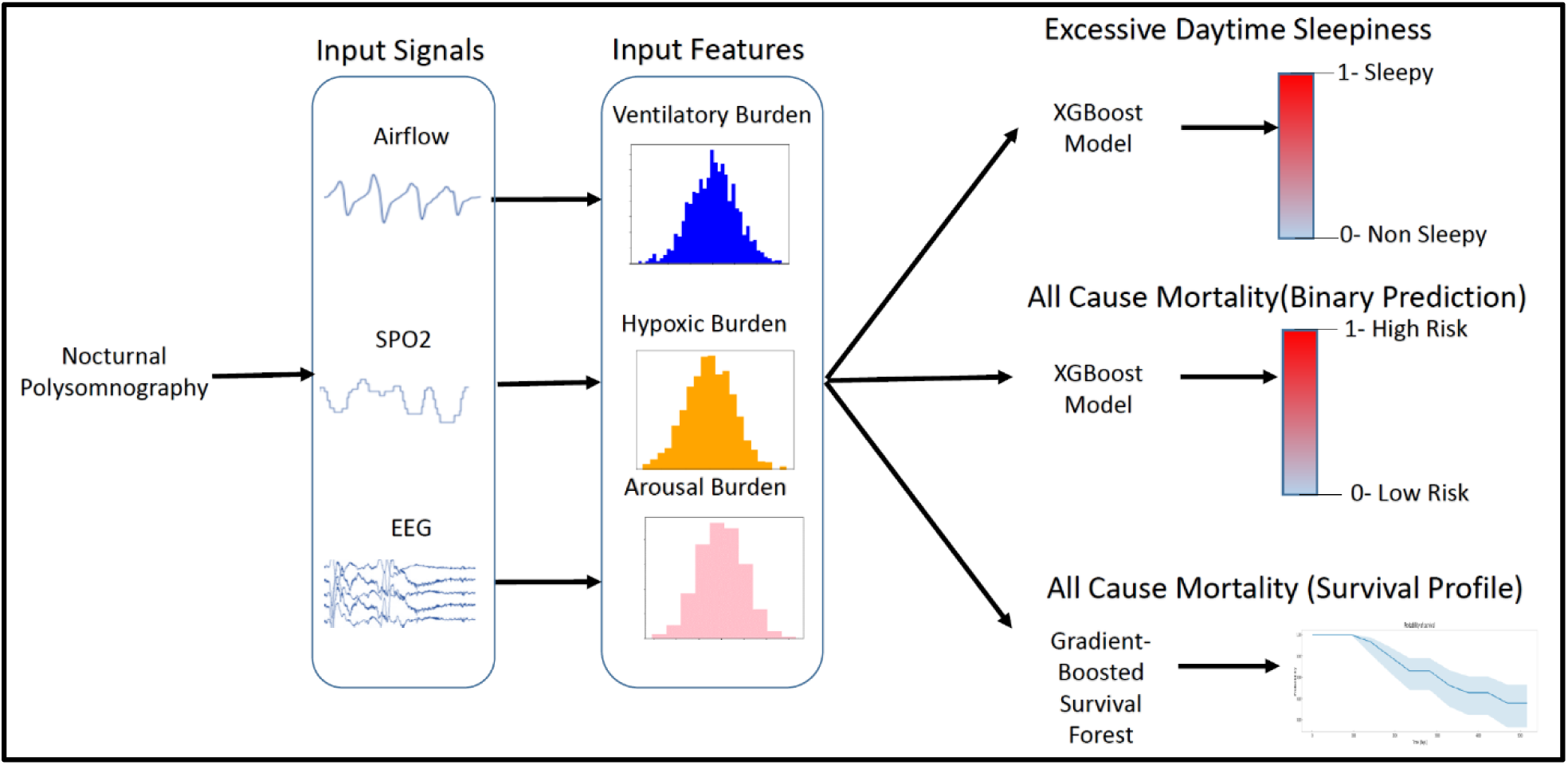
System Overview

**Figure 2:**
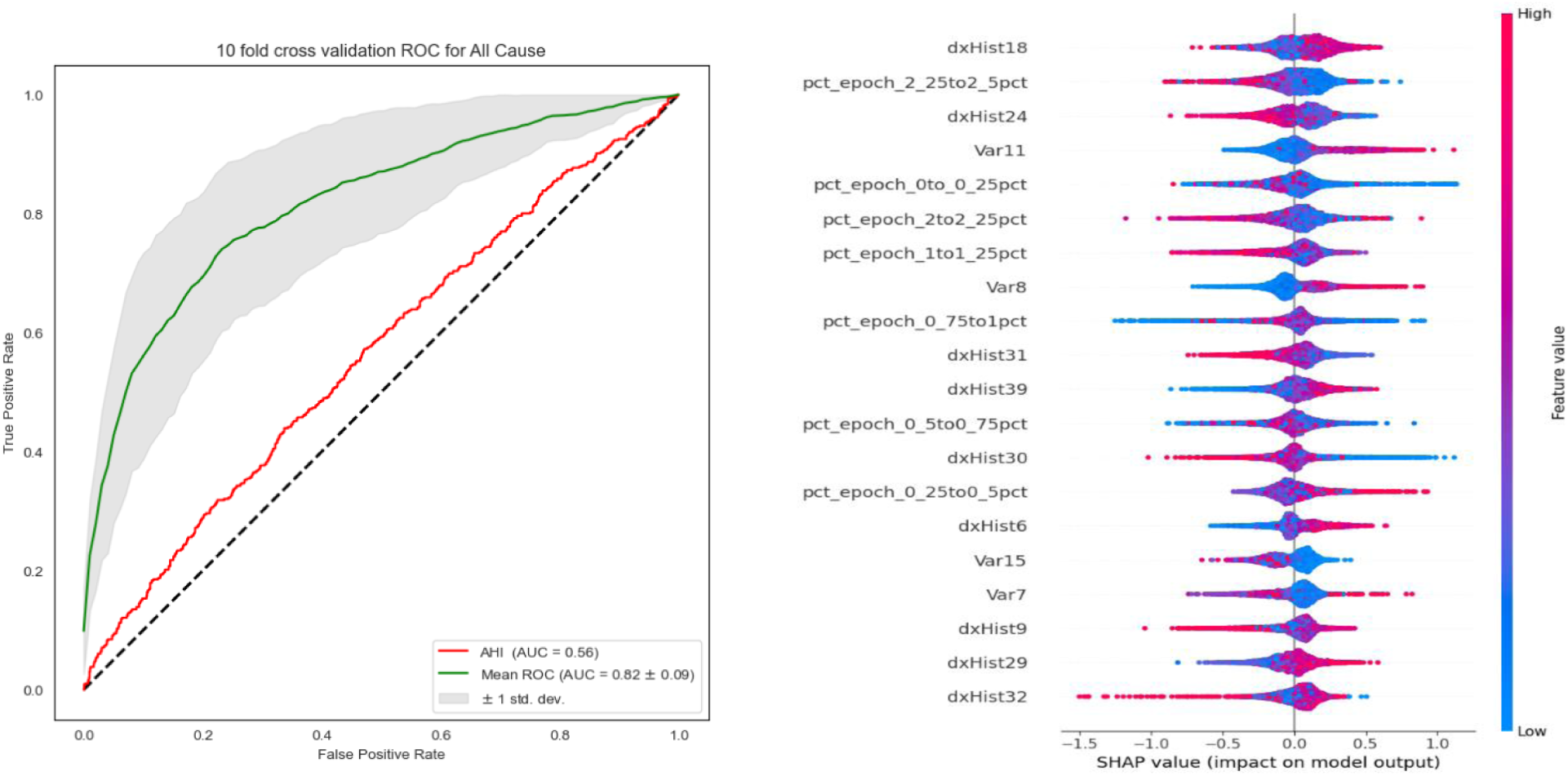
a) The AUROC curves generated for Excessive Daytime Sleepiness Prediction for ML model trained with Ventilatory, Arousal and Hypoxic Distributions and the single variable AHI model. b) SHAP feature importance generated for the ML model trained with the distributions

### Sleepiness Prediction

Fig. 4 shows the ROC curves obtained for Daytime Sleepiness using AHI and our ML model. The ML model obtained an AUROC value of (0.81±0.09), significantly better than the AHI model (logistic regression as only a single predictor is used) with an AUROC of 0.58. Fig 3 shows the most important features in predicting daytime sleepiness based on mean SHAP value. For Daytime Sleepiness we do not observe any feature representing hypoxic domain, with the majority of the features coming from the ventilatory domain.

**Figure 3:**
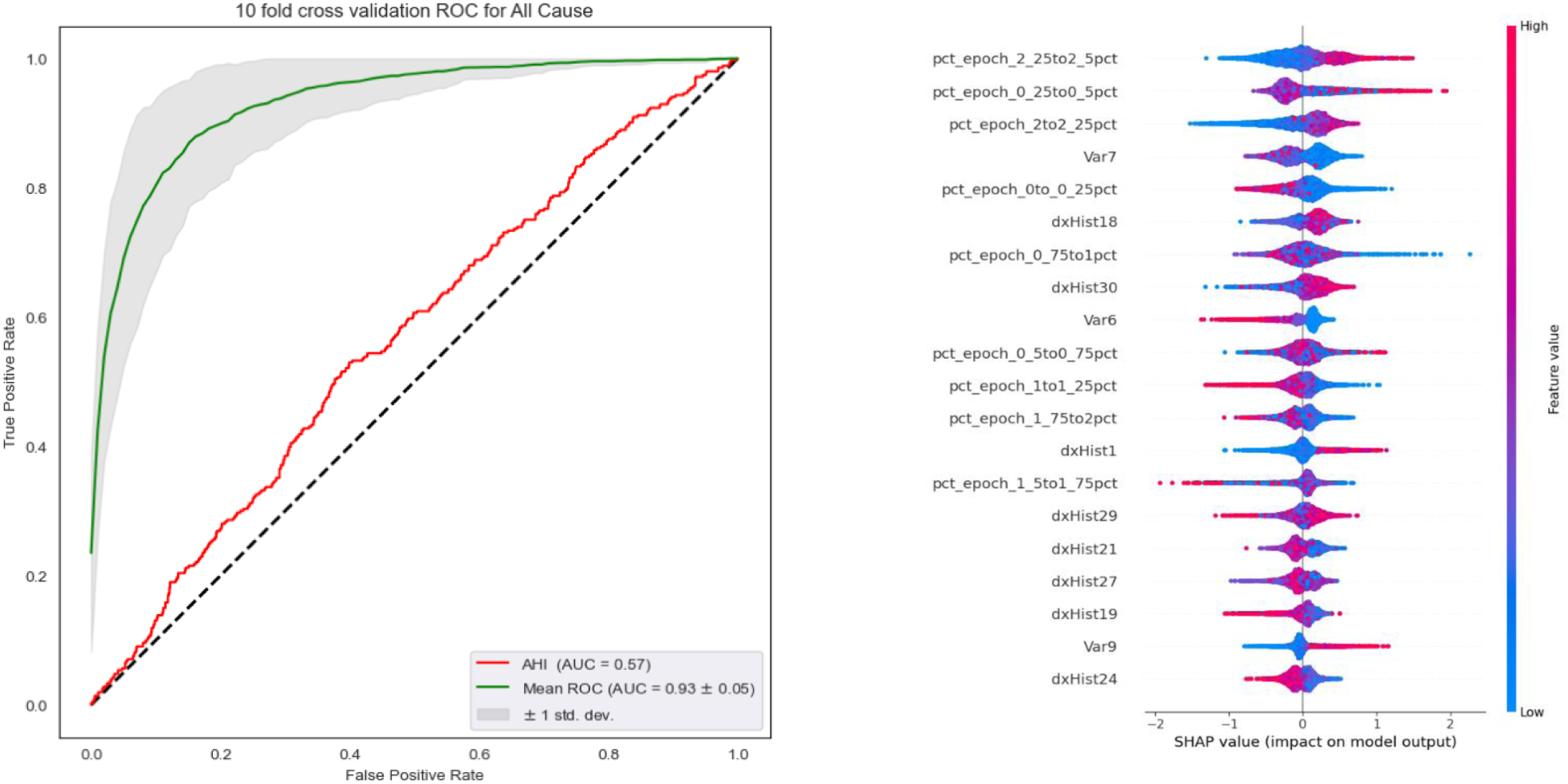
a) The AUROC curves generated for All Cause mortality Prediction for ML model trained with Ventilatory, Arousal and Hypoxic Distributions and the single variable AHI model. b) SHAP feature importance generated for the ML model trained

**Figure 4.**
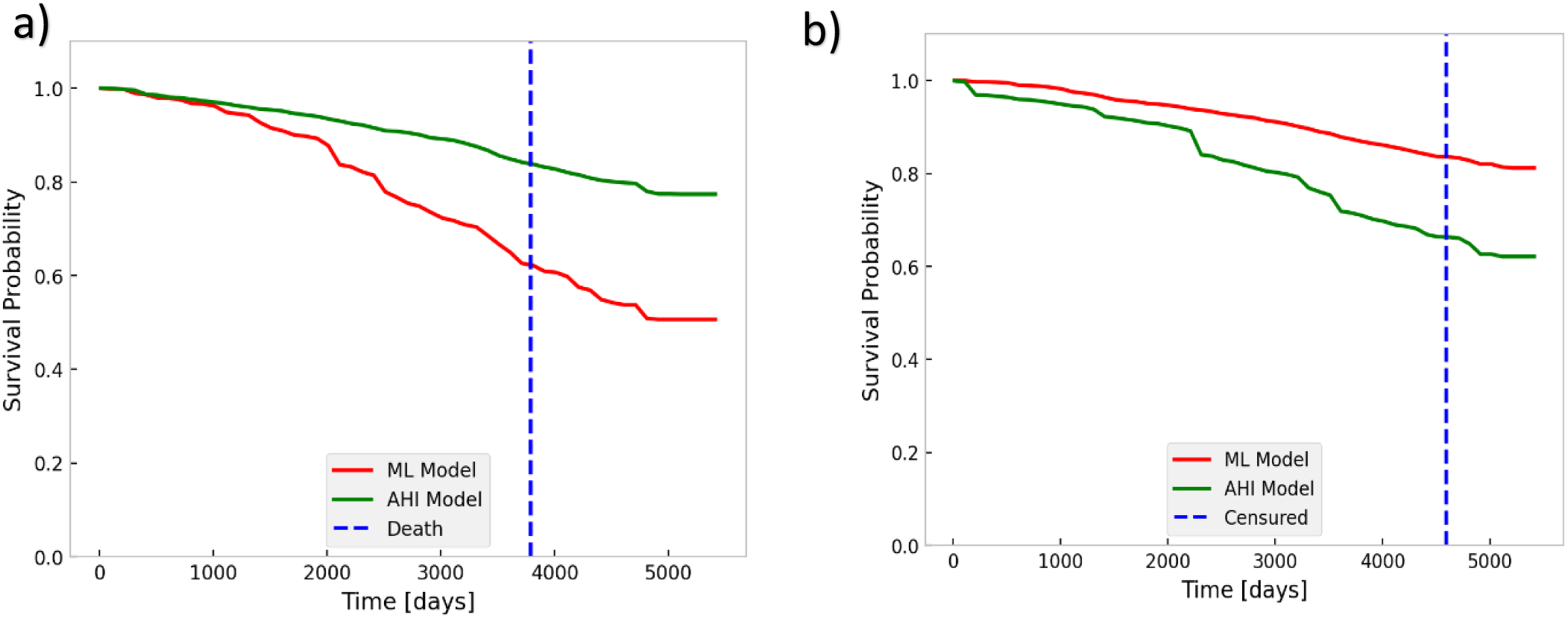
a) 1 Survival Curves generated for a subject using ML mode l(red)) and AHI model(green) for a subject who died after 3788 days Figure 4 b) Survival Curves generated for a subject using ML mode l(red)) and AHI model(green) for a subject who survived(right censored-day 4589)

### Survival Analysis

The overall performance of the survival models was evaluated using the Concordance Index (Harrell’s C-index), Integrated Brier Score and the Time dependent AUROC. Figure 5 illustrates the survival curves generated for two subjects using the proposed ML and AHI models. Fig.5 a) shows the survival curves for an individual who died during the observation period. The survival curves almost coincide with each other up to ∼1200 days. Afterward, a clear deviation was observed, with the ML model predicting a lower survival probability as time progresses than the AHI model. Fig.5 b) shows the survival curves for a right-censored individual for the two models: the ML model shows that the survival probability is higher than the AHI model.

**Figure 5:**
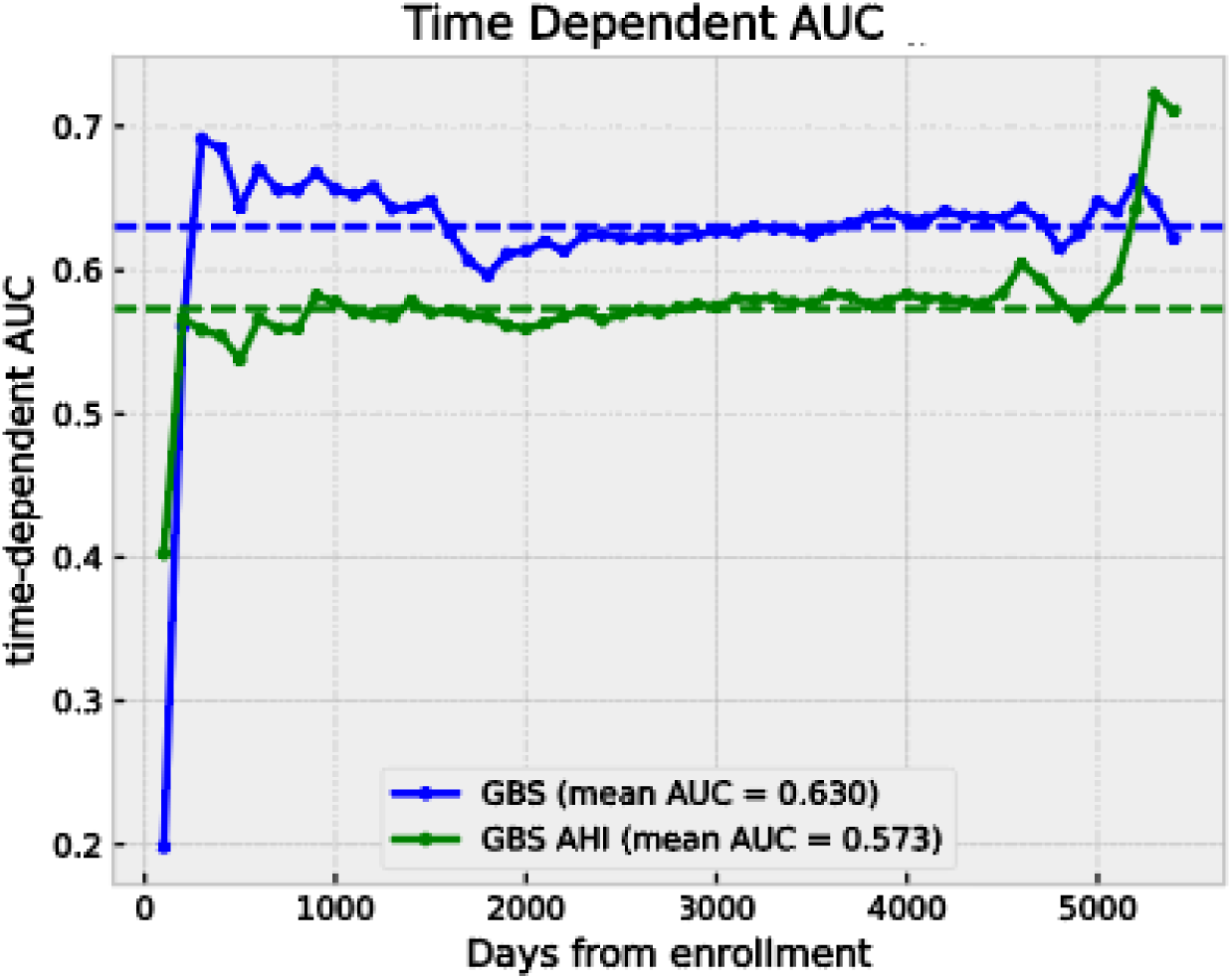
Time Dependent AUROC for the two Survival models trained with VD/AD/HD and AHI.

From the plot shown in Figure. 4 we can observe Time Dependent AUROC curves generated for the two models. The proposed ML model is performing moderately on average with an AUC of ∼0.64 (dashed line). However, there is a clear difference in performance between the first and second half of the time range. The performance on the test data drops initially and stays close to the mean value up to 4200 days (about 11 and a half years) from enrollment. After 4200 days the model performance steadily increases. Thus, we can conclude that the ML model is most effective in predicting death in towards the end of the considered time period. Throughout the time, our ML model’s performance remains better than the AHI survival model. The AHI model performs slightly better than the random model with a mean TD-AUROC of 0.57.

## Discussion

AHI has been used to clinically assess the severity of Obstructive Sleep Apnea in the field for decades. Many researchers have highlighted that AHI has been a flawed metric for decades. However, this flawed metric is still used to predict OSA’s long-term and short-term outcomes, resulting in poor predictive performance. The AHI metric combines the ventilatory, hypoxic, and arousal domains in a fixed linear combination. Previous research showed evidence that features along each of these domains are better predictors of short- and long-term outcomes of OSA than those of AHI. However, combining these domains in the optimal to improve the prediction of these outcomes is still an active research area.

For evaluating severity in individuals who are suspected of OSA, several studies have attempted the use of AI/ML methods. To categorize OSA participants into no/mild/moderate/severe OSA, Gutierrez et al. investigated the use of SpO2 data from at-home nocturnal oximetry ^30,31^. They implemented AdaBoost based ML method that can accurately distinguish between patients who experience daytime sleepiness and those who do not. There are several studies that utilized a variety of AI/ML models to detect OSA through various signals such as Airflow, ECG, and SPO2^32–34^.

In this study, we used hand-crafted features generated along each of these three domains, which can represent a whole night’s sleep. These features were used to train machine learning models, which produced a nonlinear combination of the features. Further, ML methods provide the tools to optimize our predictive models for each outcome, allowing us to build a specialized model for each outcome, unlike having a fixed combination like AHI to predict different outcomes.

In this study, we developed machine-learning models for predicting all-cause mortality and excessive daytime sleepiness through training and testing using features extracted from 3140 polysomnography records from SHHS. We have demonstrated that machine-learning models, which have thus far primarily been explored as screening or diagnosis tools in the medical field, have substantial utility in the prediction of short- and long-term consequences of OSA and generating survival profiles based on our results.

Throughout the last few decades, researchers have highlighted the importance of an alternative metric that can better capture the consequences of OSA compared to AHI. With the advancements in computational power and access to large datasets, ML-based approaches have been adopted in many medical applications to predict outcomes. Our results suggest that an ML model that represents a nonlinear combination of OSA measures across Ventilatory, Hypoxic, and Arousal domains better predicts short-term and long-term consequences of OSA compared to AHI, the traditional metric. The proposed explainable ML approach is a step towards establishing a new metric to accurately predict the outcomes of a heterogeneous disease such as OSA. Based on the feature importance calculated through SHAP analysis, it is clear that even though the same feature set was used to train the two models, the nonlinear combinations of the two outcomes differ. Hence, using a fixed combination of the three domains, such as AHI, to predict outcomes may lead to inaccurate results. Results from classification and survival curve generation show that the ML model trained with our hand-crafted features outperforms the model trained with AHI, which is the industry standard.

AI models are often considered black boxes, making interpreting the reasoning behind their predictions challenging. This lack of explainability can be a concern in healthcare, where clinicians must understand the basis before making a final decision. This study used explainable AI methods such as SHAP analysis and permutation, essential to interpret the model and innovatively use clinically relevant features. However, validating our model and model interpretations with diverse data cohorts is essential before clinical implementation. By combining the capabilities of current ML techniques with the explainable AI, our goal was to develop a model that was accurate and transparent and could be implemented clinically in the future. Our current prediction models do not utilize demographical information (age, sex, BMI, etc.) and are purely based on the PSG expression of abnormal sleep and breathing patterns. Our ML-based method has the potential to be easily implemented in a PSG processing pipeline and to be reported along with the standard sleep reports, making it accessible for clinical decision-making.

## Limitations

This paper presents a new approach that combines knowledge from physiology and the advancements of Machine Learning to predict consequences of OSA. However there are some limitations for the deployment of this model in clinical settings. These models were developed with data from epidemiological cohort and lacks clinical data. Further the models were developed using an unbalanced dataset. Although techniques were implemented to rectify issues widely associated with unbalanced data, still it cannot replace good quality data. Through SHAP analysis we provided and interpretation of the model, this analysis is only valid for this study.

## Conclusion

In this study, we aimed to combine ML methods, Explainable AI, and clinical knowledge to develop a metric that can be interpretable, accurate, and clinically implemented to determine OSA’s short- and long-term outcomes. Our analysis using a well-characterized cohort (SHHS) showed that an ML model trained with features extracted along ventilatory, hypoxic, and arousal domains may better predict OSA’s consequences than AHI. Further, the survival analysis showed that the gradient-boosted tree model trained with the features from three domains outperforms the based survival model. As a future direction, we aim to validate our findings with more data cohorts and subjects from clinical trials. In the future, we aim to assess whether adding demographic information improves the performance of our ML models.

## Data Availability

The dataset used in this study was obtained exclusively from the Sleep Heart Health Study (SHHS) hosted on the National Sleep Research Resource (NSRR) platform. This dataset is publicly available and can be accessed at https://sleepdata.org/.

https://sleepdata.org/datasets/shhs

## Supplementary Material

### Sleep Heart Health Study (SHHS)

The SHHS was a multi-center community-based prospective study designed to examine CVD consequences of sleep-disordered breathing. Details of the study design have been reported previously.^35^ A total of 6,441 men and women aged 40 years and older completed the baseline examination (1994 – 1995) which consisted of a Sleep Habits Questionnaire, anthropometric information, and overnight at-home unattended polysomnography. IRB approval was obtained for all participating institutions and all participants signed informed consent. Of the 6,441 participants, 637 participants withdrew their consent due to sovereignty issues, and as a result the final dataset comprised 5,804 subjects. The flow of subjects for the present analyses is listed in Table S1. Pre-requisites for present analyses were good quality PSG data including the abdominal and thoracic respiratory inductance plethysmography (RIP) belts. All PSG studies were reviewed manually by the study team and coded from a scale of 1 (bad) to 4 (good) for quality of PSG signals. Further, the duration of good quality signals was also denoted. Only studies that had both thoracic and abdominal RIP and SpO2 signals of good quality (scale rating 3 or 4) for more than 4 hours were used. A total of 657 participants did not have data for the covariates and hence the final number of subjects with both good quality data and covariates was N=4784. Out of these subjects, 629 did not have data on CVD mortality and hence for Cox models with the primary outcome of CVD mortality had only N=4155 subjects.

### Automated Ventilatory Distribution (VD)

#### Automated Hypoxic Distribution (HD)

Automated Hypoxic burden was defined as the total area under the desaturation curve associated with >=3% desaturation events that were identified automatically. In the present analyses, we used the hypoxic burden calculation. Prior to calculation of the total area under the desaturation curve, the SpO2 signal was pre-processed as follows. Periods of wake identified using manually scored sleep/wake were discarded. Artifacts that indicated non-physiological values were also discarded (e.g., values outside 56-100% oxygen saturation). A Savitzky-Golay filter was then applied to the discontinuous segments to create an interpolated and continuous SpO2 signal. SpO2 nadirs were identified automatically for this pre-processed signal using a peak-prominence value of 3%, i.e., decreases in SpO2 by more than 3% over 3 seconds or more were deemed as a candidate “event”. The left- and right-peak associated with these candidate events were identified using the second derivative of the pre-processed SpO2 signal. The area bounded by these peaks and the identified nadir was then evaluated and the cumulative sum of such desaturation events was then defined as the hypoxic burden

#### Arousal Distribution (AD)

Represented by Delta SWAK histogram## write the **Demographic Information-** The demographic variables included in this analysis were age, BMI and gender which were available with the SHHS data sheet.

#### Other Covariates-

##### Model Information

Excessive Daytime Sleepiness: XGBoost Model Parameters

~~~
objective=‘binary:logistic’,
eval_metric=‘auc’,
colsample_bytree=‘1.0’,
learning_rate=0.1,
max_depth= 12,
n_estimators= 300,
scale_pos_weight=9,
subsample= 0.8,
reg_alpha= 0.4,
reg_lambda=0.8,
booster="dart"
~~~

